# Using big data analytics to explore the relationship between government stringency and preventative social behaviour during the COVID-19 pandemic in the United Kingdom

**DOI:** 10.1101/2021.07.09.21260246

**Authors:** Noor Al-Zubaidy, Roberto Crespo, Sarah Jones, Reza Drikvandi, Lisa Gould, Melanie Leis, Hendramoorty Maheswaran, Ana Luisa Neves, Ara Darzi

## Abstract

We evaluated the association between preventative social behaviour and government stringency. Additionally, we sought to evaluate the influence of additional factors including time, need to protect others (using the reported number of COVID-19 deaths as a surrogate measure) and reported confidence in government handling of the COVID-19 pandemic. We used repeated national cross-sectional surveys the UK over the course of 41 weeks from 1st April 2020 to January 28th, 2021, including a total of 38,092 participants. Preventative social behaviour and government stringency index scores were significantly associated on linear regression analyses (R2 =0.6468, p<0.001, and remained significant after controlling for the effect of reported COVID-19 deaths, confidence in government handling of the pandemic, and time (R2=0.898, p<0.001). Longitudinal data suggest that government stringency is an effective tool in promoting preventative social behaviour in the fight against COVID-19.

## Introduction

Preventative social behaviours (PSB) including school and workplace closures, restrictions on mass gatherings and lockdown measures are effective in reducing transmission of COVID-19. ^1-3^ Timely implementation of these measures has been essential to control the COVID-19 pandemic that has already claimed 3.5 million lives worldwide (approximately 128,000 in the UK). ^3, 4^ Their success depends on a multitude of factors that affect the public’s adherence ^5^. A key motivating factor for adherence to PSB guidance is the individuals perceived need to protect themselves, their relatives and their community networks. ^6, 7^ The reported COVID-19 deaths have previously been used as a surrogate measure representing the need to protect oneself or family. ^8^ Major barriers to adherence to PSB guidance include lack of trust in the prescribed guidance, failure of others in compliance to the guidance, the inability to follow it (e.g., need to attend work or carer for others) and negative impact on ones’ mental health. ^8^ Additionally, lower levels of trust in governments’ public health messaging was associated with poorer adherence to PSB. ^9^

Government policy and regulations play a prominent role in the effective implementation of public health measures to tackle the COVID-19 pandemic. ^2^ The level of stringency derived from the enacted government policy has been shown to affect the levels of COVID-19 associated deaths. ^10^ A lower degree of Government stringency and slower implementation of guidance response times have been found to be associated with a higher COVID-19 related mortality. ^3^ Government policy is thought to be fundamental in promoting adherence to PSB. In the United Kingdom (UK), the government first introduced PSB measures on 16th March 2020, advising the public to avoid unnecessary travel and to work from home when possible. ^11^ One week later a nationwide lockdown was introduced, lasting for 7 weeks. Following the initial lockdown, the level of government stringency (GS) varied greatly, until a second lockdown was further introduced on November 5th, 2020, ^12^ and a third one on January 6^th^, 2021. ^13^ There is scarce evidence on how to promote adherence to PSB, specifically evaluating the relationship between governmental stringency and adherence to these guidelines over time.

Governments worldwide acknowledge that “lockdowns” are not sustainable in the long-term, it is important to understand how governmental guidance, and specifically how government stringency, has impacted the promotion of PSB by the public. ^14^ The main objective of this work was to evaluate the association between PSB and GS over time. Additionally, we sought to evaluate the influence of additional factors on the relationship between GS and PSB, including time, need to protect others (using the reported number of COVID-19 deaths as a surrogate measure) and reported confidence in government handling of the COVID-19 pandemic.

## Methods

### Study overview

Since the pandemic began, YouGov has been commissioned to undertake repeat national cross-sectional surveys the UK. In this study we examined the data collected from the UK population. This data is collected from a nationally representative cross-section of the UK population over the course of 41 weeks from 1st April 2020 to January 28th, 2021. The duration of this study covers the first national lockdown, ending on 1st June 2020, easing of restrictions during the summer and the reinstatement of the national lockdown on the 6th of January 2021. The responses to the questions in the YouGov survey were used to generate a novel preventative and social index (PSB) score and a Confidence in Government Handling index (CGH). Data from the Oxford COVID-19 Government Response Tracker (OxCGRT) was used to generate a Government Stringency (GS) index. ^15^ Ethical approval was waived as part of the Imperial College’s ethical policy.

### Participants and procedures

For each country and survey, YouGov samples a nationally representative population from a larger panel of respondents in each country to complete an online survey. YouGov generates survey weights to adjust the samples to be nationally representative. The surveys examine the populations attitudes to, and adoption of preventative social behaviours around COVID-19 and are repeated every one to two weeks (median time between survey waves of 8 days, range 5-20 days). The survey consists of a core set of demographic questions and additional questions relating to COVID-19 and preventative social behaviours to curtail the pandemic.

### Measures

In this study we used the 20 preventative social behaviour questions asked in the YouGov surveys to create a novel **PSB index**. The questions were included if they were in accordance with WHO’s COVID-19 public guidance, ^16^ if they were adopted in the last week, were in line with WHO’s recommendations, and had generalised applicability. The questions were excluded if they related to behaviours that did not represent WHO recommendation (e.g., eaten separately at home); a hypothetical question (e.g., how likely are you to self-isolate if you have symptoms); common behaviour which predates the pandemic (e.g., covering of mouth when sneezing); a behaviour which was not possible/accessible over the entire course of the pandemic (e.g., use of hand sanitizer); in contrast with government recommendations; necessary for survival; or if it may be interpreted as ambiguous (e.g., avoided going to the shops).

After reviewing the questions asked in the YouGov surveys against the inclusion and exclusion criteria, seven preventative behaviour questions were included in the PSB index. These were: (1) avoiding public transport; (2) avoiding crowded areas; (3) avoiding large social gatherings (more than 10 people); (4) avoiding medium sized social gatherings (3-10 people); (5) avoiding having guest over; (6) avoided going out in general; and (7) avoiding contact with people who have exhibited symptoms or been exposed to COVID-19. Table 1 shows the complete list of all the preventative social behaviour questions asked in the survey, and the final ones included the PSB index.

**Table 1:** Preventative social behaviours (PSB) assessed for inclusion or exclusion in the PSB index.

| **Question: Thinking about the last 7 days, how often have you taken the following measures to protect yourself or others from coronavirus (COVID-19)? As a reminder, please exclude any measures that you have already taken for reason other than coronavirus (COVID-19)** | **Inclusion in the PSB index** |
| --- | --- |
| Avoided contact with people who have symptoms or who I think may have been exposed to the virus | Included |
| Worn a face mask outside | Excluded |
| Avoiding large social gatherings (10 people) | Included |
| Avoided medium social gatherings (3-10 people) | Included |
| Avoided having guests over | Included |
| Avoided crowded areas | Included |
| Avoided going out in general | Included |
| Avoided public transport | Included |
| Slept in separate rooms from | Excluded |
| Eaten separately at home | Excluded |
| Avoiding letting children going to school/university | Excluded |
| Cleaned frequently touched surfaces in the home | Excluded |
| Washed hands with soap and water | Excluded |
| Used hand sanitiser | Excluded |
| Avoided touching objects in public | Excluded |
| Avoided small social gatherings (2 people) | Excluded |
| Avoided going to hospital or other healthcare settings | Excluded |
| Covered nose or mouth when sneezing or coughing | Excluded |
| Avoid working outside your home | Excluded |
| Avoided going to shops | Excluded |

Each of the included preventative questions from the YouGov survey, had five possible responses: ‘always’; ‘frequently’; ‘sometimes’; ‘rarely’; or ‘not at all’. Each response was ranked from most preventative (‘always’ as 5) to least preventative (‘not at all’ as 1). ^16, 17^ A weighted average of the sum of all the responses for each question for each week was calculated by multiplying the proportion of responses to the respective ranks while taking into consideration the survey weights of each participant. The mean weighted average of each of the relevant survey questions has then informed the calculation of the PSB composite index score for a specific week (PSB index weekly score = μ(*x*_1_+*x*_2_+*x*_3_+….+*x*_7_)). The PSB index values range between 1 and 5, where 5 would represent the most preventative social behaviour and 1, the least preventative.

Confidence in Government Handling (CGH) of the COVID-19 response was asked of all participants in the YouGov COVID-19 surveys. The question asked was ‘how well do you feel the government is handling the issue of coronavirus?’. The percentage of individuals responding ‘very’ or ‘somewhat well’ for each of the corresponding weeks of the published preventative behaviour survey questions was calculated to define the CGH index. ^18^

The Government Stringency (GS) index was calculated using data from the Oxford COVID-19 Government Response Tracker (OxCGRT).^15^ The OxCGRT systematically collects information on several different common policy responses that governments have taken to respond to the pandemic on 20 indicators such as school closures and travel restrictions for more than 180 countries to form different indices, one of which is the ‘stringency index’. The ‘stringency index’ records the variations in responses from publicly available information for each day, which is calculated as a composite measure based on nine indicators measured on an ordinal scale (from 0-100, where 100 is the most stringent). The nine indicators used are: school closures, workplace closures, cancelled public events, restrictions on gatherings, closure of public transport, public information campaigns, stay at home measures, restrictions on internal movement and international travel controls.^19^ As each wave of questionnaires was sent over a period of 2-4 days, the mean of the highest and lowest stringency index score was calculated to form the GS index for the period over which the waves were delivered.

The 7-day average of the number of deaths of people who had had a positive test result for COVID-19 and died within 28 days of the first positive test (Reported COVID-19 deaths, RCD), as reported by Public Health England (PHE) was recorded for each of the corresponding weeks of the published preventative behaviour survey questions.^4^

### Data Analysis

The characteristics of participants including age, gender and geographical location were based on responses to questions asked in the survey. We examined at the relationship between PSB index scores, GS index, CGH, and recorded COVID-19 deaths between April 2020 – April 2021, across four significant events, including the start of the 1st national lockdown (April 2020), the launch if the ‘Eat out to help out’ government initiative (August 2020), the start of the second lockdown (November 2020), and the start of the third lockdown January 2021). We explored how changes in GS was related to the adoption or PSB (measured by our PSB index), for this we used the GS score on the date of the first lockdown, and corresponding PSB score as a reference score and compared the % change over the following surveys. We next explored whether the relationship remained significant after controlling for the effect of reported COVID-19 deaths (as a proxy to protect others), CGH and time (as well as potential interactions between GS index and time, and between CGH and time).

Data was initially visualized using in Excel (v 16.49), where line and scatter plots were created to explore the data. Statistical analysis was conducted using RStudio Desktop (v1.4.1103), and R (v 4.0.5). Linear regression analysis, using the ordinary least squares method, was conducted using the using base R ‘lm’ function, and R^2^ and p values were extracted using the ‘summary’ function. The association of the PSB index (described below) with the variables of interest was determined first using simple linear regressions, using the PSB index as the outcome variable. Further analyses were conducted using multiple linear regression to determine the influence of variables, such as time, CGH and RCD and GS index on the association between the PSB index.

## Results

Over the 41 weeks considered in this study, there were 38,092 participants answering the 7 questions related to the PSB index in the UK, across 37 survey waves that took place in between 01/04/2020, and 25/03/2021. Survey participants were aged between 18 and 70 years and represent the different regions of the UK. A more detailed overview of the participants’ characteristics is provided in **Table 2**.

**Table 2:**
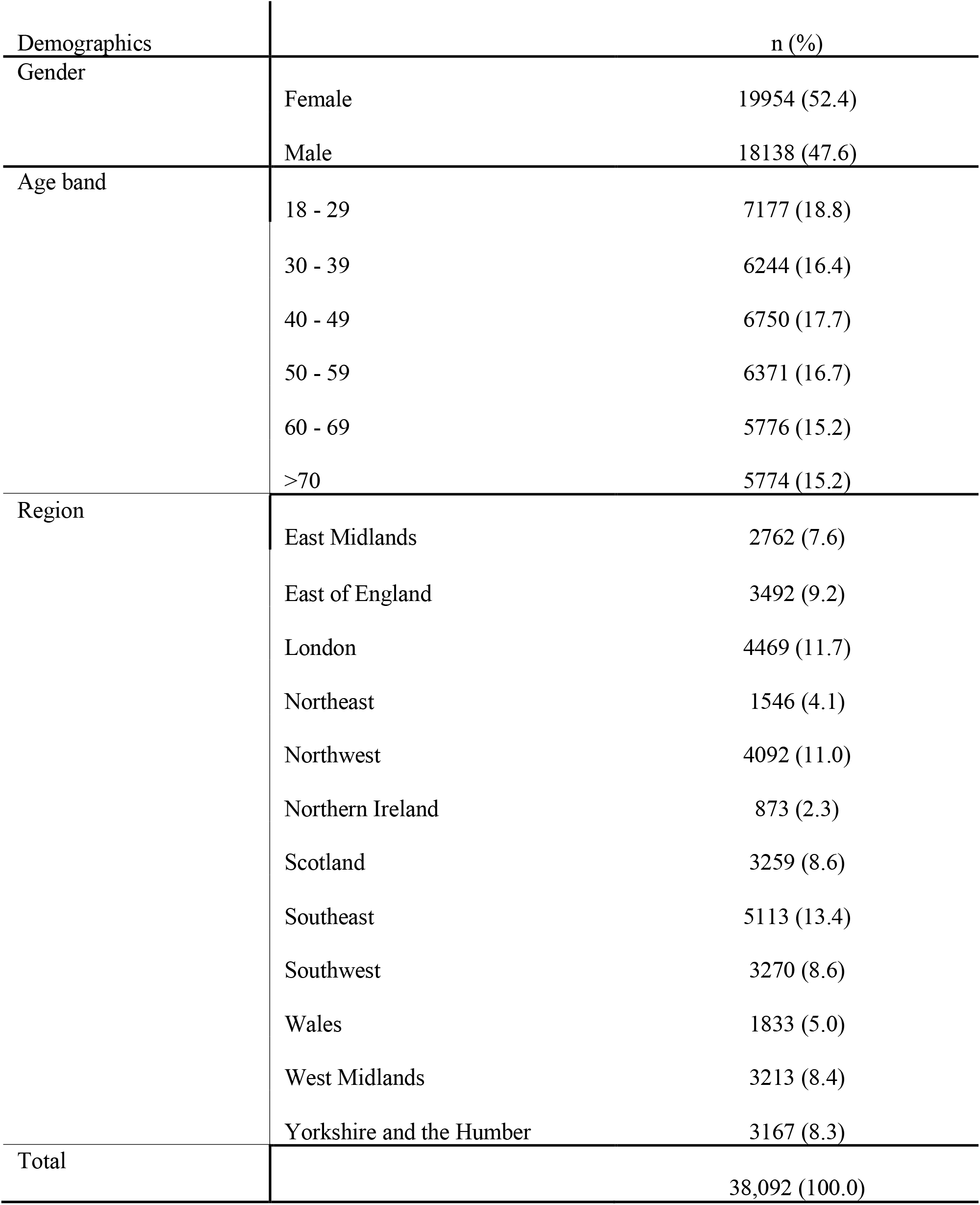
Participants’ characteristics.

### Association between preventative social behaviour and government stringency

The highest PSB index score (PSB=4.68) was observed during the first national lockdown (April-June 2020. The corresponding GS index score was 79.60. During the first lockdown the PSB index score ranged between 4.39-4.68 and the GS index score ranged between 70.35-79.60. During the second short lockdown, the PSB index score was 4.249 and the GS 67.59. During the third lockdown the PSB index score ranged between 4.350-4.580 and the GS index score ranged between 78.70-87.96. A detailed overview of the variation of PSB and GS indices, as well as recorded COVID-19 deaths is provided in Table 3.

**Table 3.** Variation of Average Preventative Social Behaviour (PSB) index, Recoded COVID-19 deaths, Government Stringency (GS) index and Confidence in Government Handling (CGH) across the duration of the study.

| Event | Survey date | PSB index | Recorded COVID-19<br>deaths (7-day<br>average) | GS index | CGH |
| --- | --- | --- | --- | --- | --- |
| 1st lockdown | 01/04/2020 | 4.661 | 724.00 | 79.60 | 72 |
|  | 07/04/2020 | 4.677 | 960.00 | 79.60 | 64 |
|  | 15/04/2020 | 4.656 | 863.10 | 79.60 | 68 |
|  | 21/04/2020 | 4.656 | 738.30 | 79.60 | 60 |
|  | 01/05/2020 | 4.583 | 524.10 | 79.60 | 58 |
|  | 07/05/2020 | 4.601 | 424.70 | 79.60 | 53 |
|  | 15/05/2020 | 4.539 | 307.60 | 71.30 | 47 |
|  | 22/05/2020 | 4.477 | 230.40 | 71.30 | 45 |
|  | 27/05/2020 | 4.391 | 195.70 | 70.35 | 41 |
|  | 02/06/2020 | 4.451 | 143.30 | 67.60 | 39 |
|  | 09/06/2020 | 4.351 | 102.60 | 73.20 | 40 |
|  | 17/06/2020 | 4.363 | 67.40 | 71.30 | 43 |
|  | 25/06/2020 | 4.104 | 57.60 | 71.30 | 44 |
|  | 03/07/2020 | 4.113 | 34.40 | 67.80 | 42 |
|  | 08/07/2020 | 3.986 | 27.60 | 64.40 | 43 |
|  | 22/07/2020 | 3.863 | 14.40 | 64.40 | 45 |
|  | 29/07/2020 | 3.859 | 12.30 | 66.90 | 43 |
| Eat out to help out<br>scheme introduced | 06/08/2020 | 3.856 | 11.10 | 69.90 | 40 |
|  | 12/08/2020 | 3.793 | 9.40 | 68.05 | 38 |
|  | 19/08/2020 | 3.717 | 7.30 | 66.20 | 40 |
|  | 27/08/2020 | 3.731 | 8.30 | 66.20 | 39 |
|  | 02/09/2020 | 3.749 | 8.10 | 64.40 | 38 |
|  | 09/09/2020 | 3.744 | 12.10 | 65.05 | 37 |
| 'Rule of six'<br>introduced | 16/09/2020 | 3.739 | 21.40 | 66.70 | 30 |
|  | 23/09/2020 | 3.837 | 37.00 | 66.65 | 30 |
|  | 03/10/2020 | 3.929 | 65.30 | 67.60 | 31 |
|  | 14/10/2020 | 3.920 | 133.30 | 62.97 | 32 |
|  | 28/10/2020 | 3.999 | 291.40 | 75.00 | 32 |
| 2nd lockdown | 11/11/2020 | 4.249 | 421.40 | 67.59 | 34 |
|  | 16/12/2020 | 4.230 | 474.30 | 71.30 | 36 |
|  | 30/12/2020 | 4.437 | 704.60 | 81.02 | 31 |
| 3rd lockdown | 13/01/2021 | 4.526 | 1194.50 | 87.96 | 37 |
|  | 28/01/2021 | 4.580 | 1039.40 | 87.96 | 34 |
|  | 10/02/2021 | 4.560 | 580.70 | 87.96 | 39 |
|  | 24/02/2021 | 4.481 | 265.30 | 86.11 | 43 |
|  | 10/03/2021 | 4.410 | 119.10 | 82.41 | 50 |
|  | 25/03/2021 | 4.350 | 40.60 | 78.70 | 49 |

Values are shown per week where a survey was conducted. Relevant events are mentioned under the 'Event' column, and the associated duration is highlighted by horizontal grey bars.

For a better visualisation of how the GS and PSB index scores have changed over time in comparison to when PSB was at its highest (on the week of the 7^th^ April 2020), PSB and GS index scores during each week are shown as a percentage increase or decrease to their relative figures on the 7^th^ April 2020, **Figure 1a**. Overall, PSB and GS index scores were significantly associated on linear regression analyses (R^2^ =0.790, *P*<0.001) **(Figure 1b)**.

**Figure 1:**
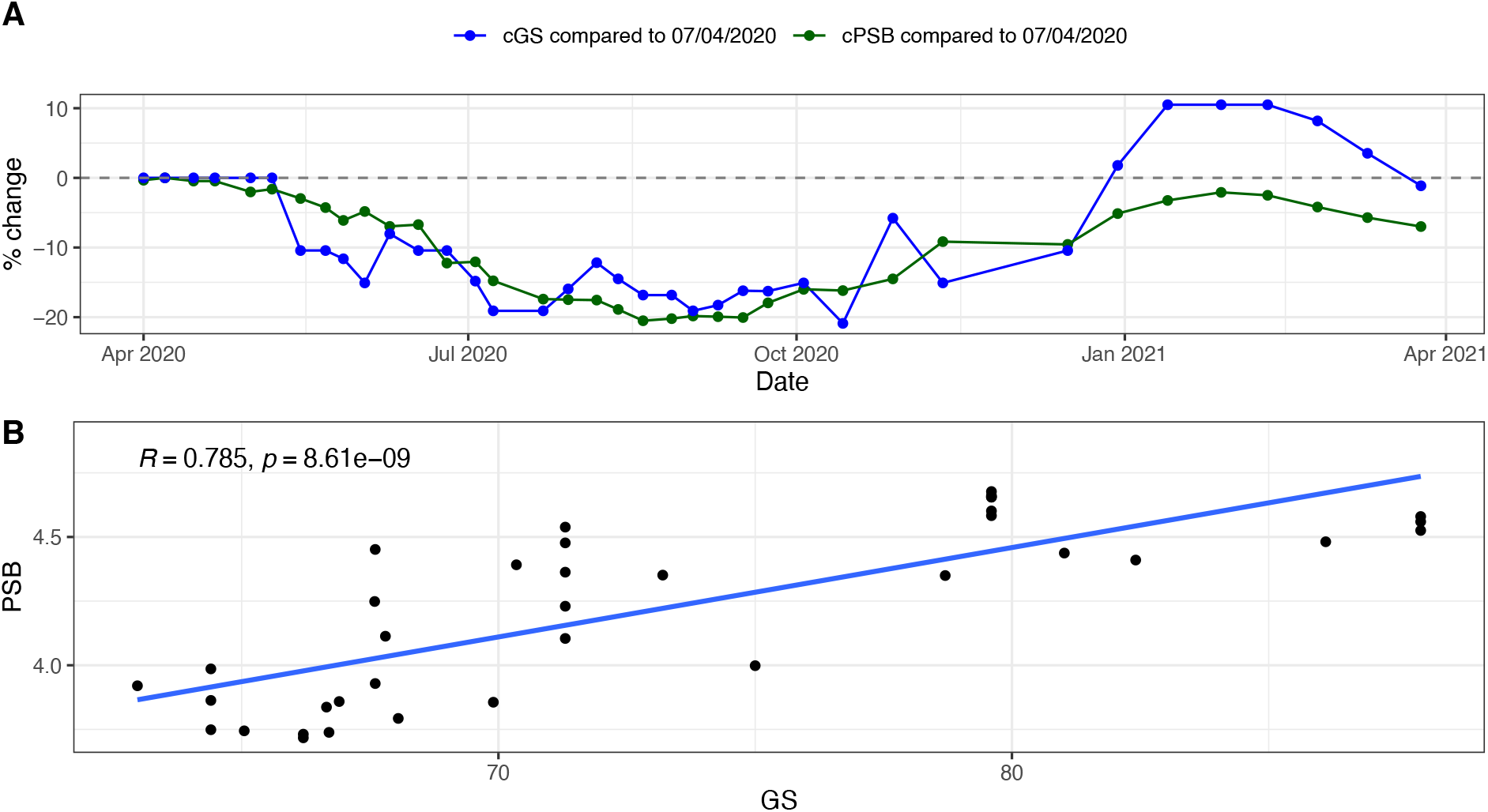
Relationship between ‘government stringency’ (GS) and ‘preventative social behaviour’ (PSB) over time. A. Percentage change in PSB and GS compared to when PSB was at its highest in the week of 7th April 2020. B. Correlation between PSB and GS (R=0.785, P < 0.001)

### Associations with time, confidence in government and COVID-19 deaths

During the first lockdown the CGH index score ranged between 39-72 and the RCD between 4143.30-960.00. During the second short lockdown, the CGH index score was 34 and the RCD 421.40. During the third lockdown the CGH index score ranged between 34-50 and the RCD between 4.350-4.580. A detailed overview of the variation across time is provided in Table 3.

Significant associations were found between PSB and RCD (R^2^=0.74, *P*< 0.001), CGH (R^2^ =0.59, *P*<0.001), and GS index (R^2^ =0.79, *P* <0.001) (**Figure 2**), but not with time. The variation of PSB index scores, GS index scores, CGH and reported COVID-19 deaths (as a proxy for the need to protect others) over time are illustrated in **Figure 3**, and weekly values shown in **Table 1**. In April 2020, PSB index had peaked at 4.68 and reported COVID-19 deaths reached an initial peak at 960 deaths per day (7-day average). During this month public opinion on government handling of the pandemic steadily declined from 72-60% over the same month. Reported COVID-19 deaths peaked again in January 2021, reaching higher levels than in April 2020, with PSB index scores increasing alongside this, but never quite reaching the same levels as in April 2020 (PSB = 4.68). All four variables continued to show a general pattern of decline over time until September 2020. PSB index scores were at their lowest in the week of the 19^th^ of August 2020, two weeks following the introduction of the ‘eat out to help out’ government scheme. This also coincided with the highest level of reported COVID-19 deaths of 1195 deaths per day (7-day average) on the week of 13^th^ January 2021. This also coincided with the highest level of GS (87.96) and the second peak of PSB (4.58).

**Figure 2:**
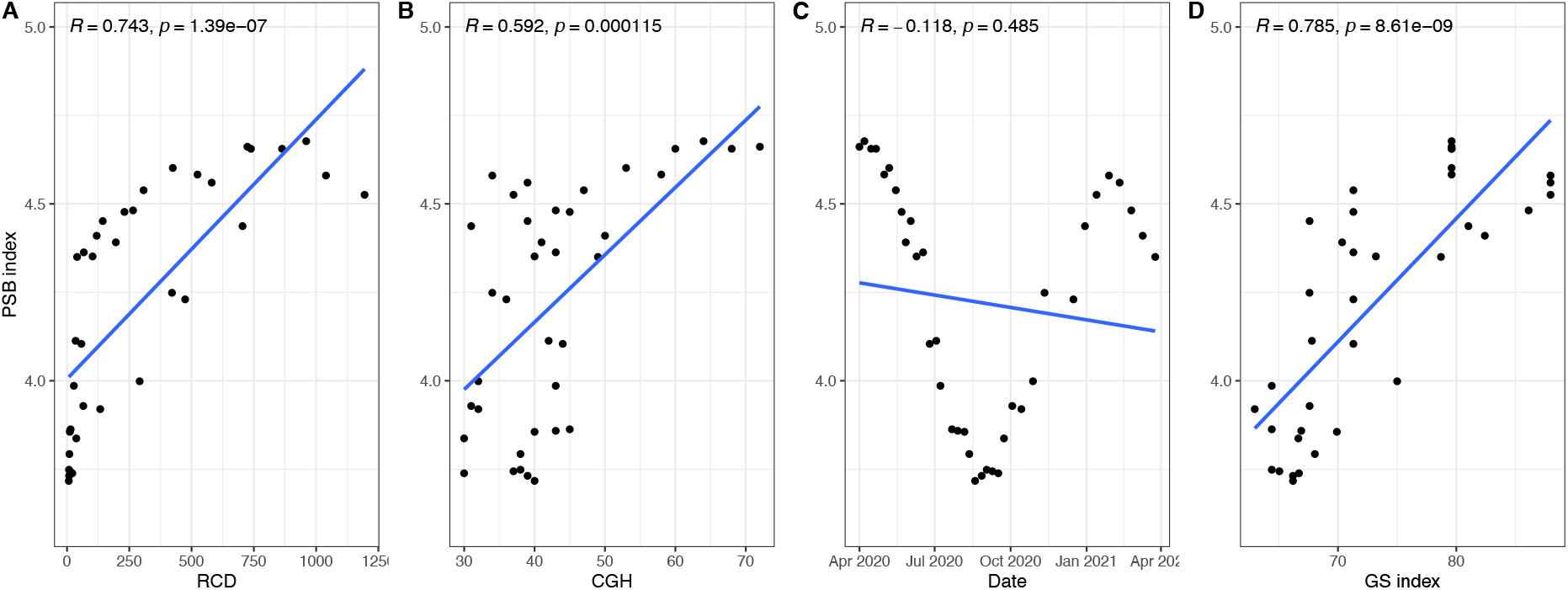
Associations between preventative social behaviour (PSB index) and recorded COVID-19 deaths (RCD), confidence in government handling (CHG), time and government stringency (GS) index. A) Relationship between PSB and RCD (7-day average) (R2=0.743, P<0.001); B) Relationship between PSB and CGH (% responding well/somewhat well, R2=0.592, P<0.001); C) Association between PSB and time (R2=0.118, P=0.485); D) Association between PSB and GS index (R2=0.785, P<0.001).

**Figure 3:**
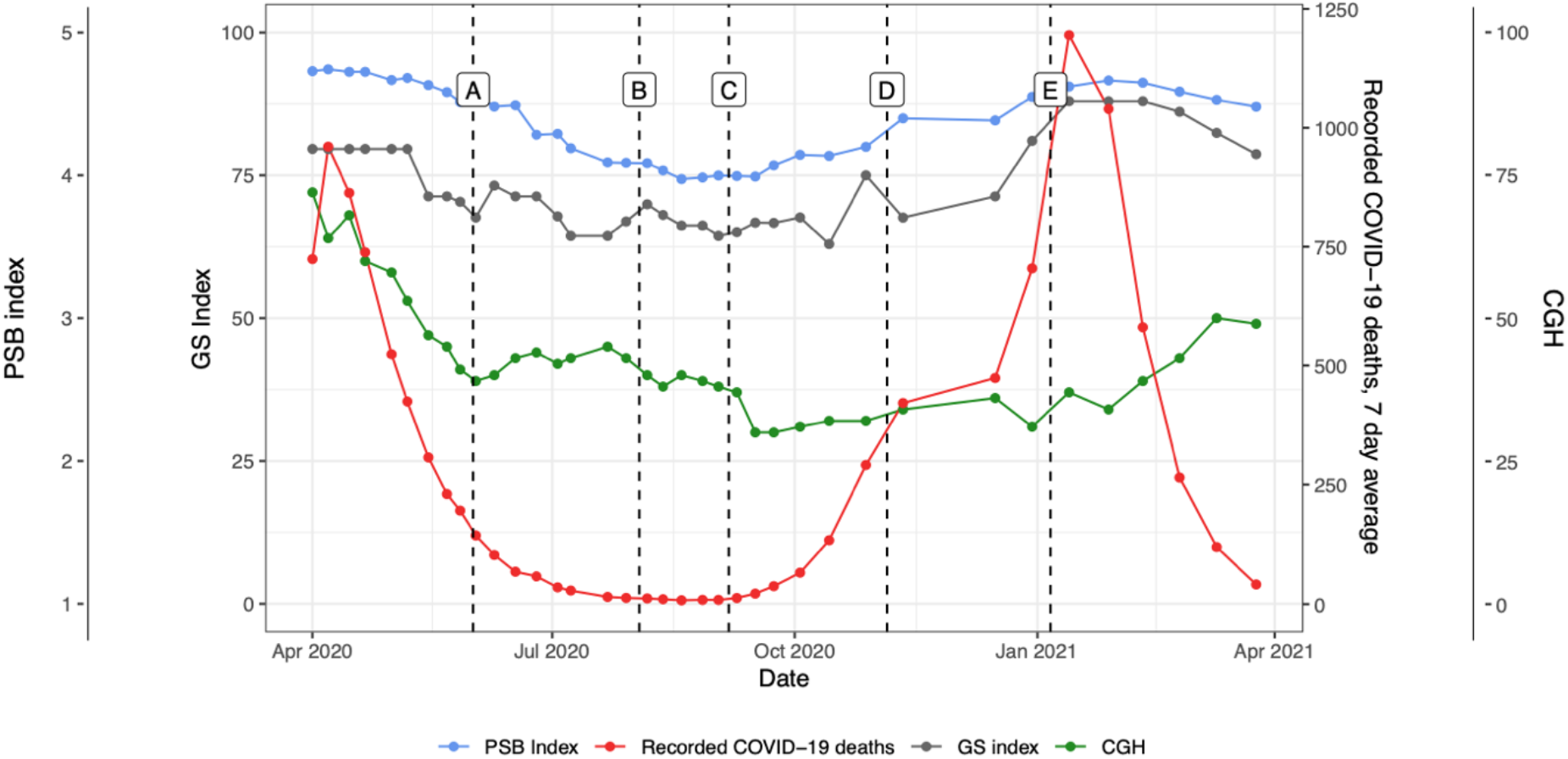
Changes of ‘PSB index’ (blue), ‘GS index’ (grey), ‘recorded COVID-19 deaths, 7-day average’ (red) and ‘reported confidence in the government’s handling of the pandemic (CGH), percentage response – handling well/very well’ (green) over ‘time’. Five important events are also represented in the graph (A-E). A: 1st June 2020, 1st national lockdown measures eased. B: 3rd August 2020, ‘Eat out to help out’ government initiative launched. C: 6-22 September 2020, R value increases to 1.2, newer restrictions on public house openings and crowds (maximum 6 allowed to gather). D: 5th November 2020, second national lockdown imposed; E) 6th January 2021, third national lockdown measures imposed.

When controlling for the effect of reported COVID-19 deaths (as a proxy to protect others), CGH and time (as well as potential interactions between GS index and time, and between CGH and time), the association between PSB and GS remained significant (R^2^=0.898, *P*<0.001). The interaction of time on GS shows a small but significant negative correlation R^2^=-0.002 (*P*=0.001), suggesting whilst an increase in GS correlates with an increase in PSB, the effect of GS in doing so may slightly reduce over time. The same effect was observed regarding the interaction of RCD and time (R^2^=0.001, *P*<0.001). Neither confidence on governmental handling (CGH), nor its interaction with time had statistical significance within the model **(Table 4)**.

**Table 4:** Multiple regression table displaying the PSB as the dependent variable, with seven independent variables.

| Dependent variable | Independent Variables | Unadjusted R2 | P | Adjusted R2 | Adjusted P |
| --- | --- | --- | --- | --- | --- |
| PSB composite index score | Intercept | 2.785 | 0.007 | 0.898 | <0.001 |
|  | GS index | 0.032 | 0.027 |  |  |
|  | Effects of 'GS index' over time | -0.001 | 0.007 |  |  |
|  | RCD | -0.000 | 0.718 |  |  |
|  | Effects of 'RCD' over time | 0.000 | 0.001 |  |  |
|  | CGH | -0.008 | 0.337 |  |  |
|  | Effects of 'CGH' over 'time | -0.001 | 0.053 |  |  |
|  | Time | 0.118 | 0.015 |  |  |

## Discussion

### Key findings

Preventative social behaviour and government stringency index scores were significantly associated on linear regression analyses. Significant associations were also found between preventative social behaviour and time, reported COVID-19 deaths (as a proxy for need to protect oneself and others), and confidence in government handling of the pandemic. The associations between preventative social behaviour and government stringency remained significant even after controlling for the effect of reported COVID-19 deaths, confidence in government handling of the pandemic and time. The interaction of time on government stringency shows a small but significant negative correlation, suggesting whilst an increase in government stringency correlates with an increase in preventative social behaviour, the effect may slightly reduce over time.

### Comparison with previous literature

Our findings add to the growing evidence around the important relationship between the degree of government stringency and the adoption and maintenance of preventative social behaviours amongst the public. ^2, 5, 6, 8, 12^ However, whilst preventative social behaviour is expected to rise with an increase in government stringency index levels, the effectiveness of government stringency on the preventative social behaviour appears to reduce over time. A reduction in the effectiveness of government stringency on preventative social behaviour over time be in keeping with current theories and findings whereby behaviour response to government recommendations is likely to flounder with passing of time. ^7^

Our results highlight the changes in the stringency of UK Government policy over time, and its negative impact on the public observing preventative social behaviours. A lack in the clarity of message may have contributed to the decline in preventative social behaviour scores as patients were unsure of which rules to follow.^11, 20^ Our results representing the percentage change in both preventative social behaviour and government stringency index scores over time, allow a more direct visual comparison between the two: in general, as government stringency scores dropped, the public was expected to relax on preventative social behaviour, as exemplified by the government initiatives to kickstart the service industry by incentivising the use, actively promoting the public to leave their homes and dine in public restaurants (i.e. ‘eat out to help out scheme’). ^21^ Therefore, a drop in preventative social behaviour in line with that of government stringency could suggest that the public is adhering aptly to government advice at both higher and lower stringency levels.

To our knowledge, no other research has followed the UK public’s behaviours over the course of the pandemic to reflect how preventative social behaviour may have altered in response to ongoing changes in stringency levels over time. The failure to reach the same preventative social behaviour index levels seen in April 2020 (4.68), in January 2021 with the highest level of stringency (87.96) may be explained by several theories. The idea of lockdown fatigue is now a popularised controversial term [19]. The rationale for the cautious and possibly delayed instigation of a national lockdown in the UK was in part due to the expected public exhaustion from lockdown and consequently eventual non-adherence to social distancing guidance, at the wrong time.^22^ Additionally, our results demonstrate the erratic changes in stringency policy in the UK, in between the two national lockdown periods, this also coincided with a drop in the PSB index. A lack in the clarity of message may have contributed to the decline in PSB scores as citizens were unsure of which rules to follow.^11, 20^ Our findings would also be in keeping with existing literature whereby a delay in implementing necessary measures were more likely associated with an increase in COVID 19 cases during the second wave, Rypdal et al’s modelling, suggested that the predicted second wave in Europe was more likely a consequence of intervention fatigue, a failure to reintroduce necessary interventions at an appropriate time. Countries which did so early were less likely to suffer the same level of fatalities, in the second wave. ^23^ Further reinforcing the idea that stringent governmental behavioural policy continues to be effective in the reduction of COVID-19 cases.

The relationship between government stringency and COVID-9 related deaths has been examined at length, ^24^ however little research has evaluated the relationship between government stringency and the underlying facets which it has been designed to alter to reduce COVID-19 related deaths. On February 14^th^, 2021, the UK had recorded over 3.84 million cases of COVID, with over 2.314 million individuals having received their first dose of the vaccine, despite this the preventative social behaviour index was as relatively high in the week of 28^th^ January as it was in May 2020. This relatively high preventative social behaviour index score may reflect several issues: the UK’s general compliance to law and rules, the lack of opportunity the public must break from adherence to recommendations and fear of punishment. ^21, 25^ As such, these theories may also provide an understanding in explaining the lack of influence public opinion may have had on the preventative social behaviour index scores within our model.

Interestingly, in May 2020, the preventative social behaviour index scores began to decline; however, the GS index scores however remained constant at 79.6 for a further two weeks. Furthermore, the PSB index managed to reach a peak at the peak of the second national lockdown, comparable to the second week of May 2020. One explanation for this may be that at the height of the first national lockdown, the effect of ‘rallying behind the flag’ was at its highest, as suggested by the level of confidence in the government’s response to the COVID-19 (60-72%). ^26^ As the confidence in the governments’ response to the COVID-19 pandemic dropped and in turn the sentiment of uniting together, the ability to achieve the same record high preventative social behaviour may have diminished.

### Limitations

The limitations of this study are those associated with an observational study using publicly available data on which our analysis has been made. As such, our findings do not infer causality, rather highlight the relationship between our examined variables and the strengths of such relationships. Furthermore, our analysis is built on several assumptions. Firstly, all behaviours within the preventative social behaviour index are weighted equally and therefore contribute equally to the level of caution we have recorded as the preventative social behaviour index. Secondly, the YouGov global survey responses are weighted by YouGov using sociodemographic data to ensure they are nationally representative. However, an average of only 1006 individuals were questioned in each wave, which may not have been an adequate sample in reliably representing an entire nation. The responses were self-reported and therefore will be allied to its associated bias. ^27^

Moreover, numerous complex variables in determining preventative social behaviour have been outlined within the literature, we however chose to review four: time, government stringency, confidence in government handling of the pandemic and reported COVID-19 deaths. It may be argued that further evaluation of specific demographics such as age, gender, employment, and mental health status may be necessary in evaluating changes in preventative social behaviour, and these should be further explored in future research.

### Implications for policy

The relationship of government stringency on COVID-19 related deaths has been examined at length, however little research has evaluated the relationship between government stringency and the underlying facets which it has been designed to alter to reduce COVID-19 related deaths, the public’s behaviour. This research allows us to appreciate a plausible relationship between changes in GS and PSB demonstrating relatively high levels of adherence to government stringency levels over time. Several theories have been postulated with regards to the efficacy of non-pharmaceutical interventions over time, however our findings would suggest that adherence to such interventions remains relatively high, even in the second wave of the pandemic in the UK and therefore the use of increasing government stringency as an intervention against the spread of COVID-19 may continue to be an effective intervention in the future should it be required.

## Conclusion

Longitudinal data has shown preventative social behaviour has typically increased and decreased with government stringency over the entire course of the pandemic. There may have been new interventions which could have improved (e.g., increase in police fines) or worsened adherence (e.g., increase in vaccinations). Despite this, there seems to be a moderate relationship between the two throughout. This may therefore suggest that government stringency is an effective tool in moderating nonpharmaceutical interventions such as promoting preventative social behaviour in the fight against COVID-19.

## Data Availability

All data for YouGov surveys are de-identified and publicly available from https://github.com/YouGov-Data/covid-19-tracker.

## Acknowledgements

We thank YouGov Plc for providing us the data. YouGov are commissioned by the UK Government to undertake the surveys.

## Contributors

RFC, SJ, MSL, HM, ALN and AD conceived and designed the study. NAZ, RFC and RD did statistical analysis. ALN wrote first draft of paper. All authors contributed to data interpretation, revision of paper and contributed to discussion.

## Data sharing

All data for YouGov surveys are deidentified and publicly available from https://github.com/YouGov-Data/covid-19-tracker.

